# Beyond Hearing Loss: Ageing as a Tinnitus Risk Factor

**DOI:** 10.1101/2023.03.02.23286668

**Authors:** L. Reisinger, F. Schmidt, K. Benz, L. Vignali, S. Rösch, M. Kronbichler, N. Weisz

## Abstract

**Background:** Tinnitus affects 10 to 15 percent of the population, but its underlying causes are not yet fully understood. Hearing loss has been established as the most important risk factor. Ageing is also known to accompany increased prevalence, however, the risk is normally seen as a consequence of (age-related) hearing loss. Whether ageing per se is a risk factor has not yet been established. We specifically focused on the effect of ageing and the relationship between age, hearing loss and tinnitus.

**Methods:** We used two samples for our analyses. The first, exploratory analyses, comprised 2249 Austrian individuals. The second included data from 16008 people, drawn from a publicly available dataset (NHANES). We used logistic regressions to investigate the effect of age on tinnitus.

**Findings:** In both samples, ageing per se was found to be a significant predictor of tinnitus. In the more decisive NHANES sample, an interaction effect was observed as well. Odds ratio analyses show that per unit increase of hearing loss the odds of reporting tinnitus is higher in older people (1.06 vs 1.03).

**Interpretation:** Expanding previous findings of hearing loss as the main risk factor for tinnitus, we established ageing as a risk factor in its own right. Underlying mechanisms remain unclear, and this work calls for urgent research efforts to link biological ageing processes, hearing loss and tinnitus. We therefore suggest a novel working hypothesis that integrates these aspects from an ageing brain viewpoint.

**Funding:** Austrian Research Promotion Agency (FFG; BRIDGE 1 project “SmartCIs,” 871232) and Land Salzburg (“Hidden Hearing Loss”, 20204-WISS/225/288/4-2021).

## Introduction

Tinnitus describes the conscious perception of a sound, such as a ringing or a hissing, in the absence of any identifiable external physical source. Most individuals will have experienced this phenomenon in a transient form at some point in their lives (e.g., following a concert). Estimates of prevalence vary, with ∼15 percent of the adult population having perceived tinnitus at least occasionally (Biswas et al., 2022), while the condition is chronic in approximately 10% of the adult population (Jarach et al., 2022). The extent to which tinnitus is experienced as bothersome varies, with approximately 3% of the population reporting at least moderate distress (Baguley et al., 2013). Thus, beyond the individual level, tinnitus creates a significant financial burden on society, i.e., in terms of healthcare costs, missed days at work or early retirement (Trochidis et al., 2021). Even though there are interventions to treat tinnitus-related distress (e.g., Hesser et al., 2011), there is currently no clinical intervention that reliably eliminates the experience of the phantom sound. This deficit on the treatment side is paralleled by an absence of reliable methods (e.g., biomarkers) to diagnose tinnitus that go beyond subjective reports. This overall unsatisfactory state is ultimately due to an insufficient mechanistic understanding of why tinnitus develops and how it is manifested in changes to brain structure and of function.

Among risk factors known to be associated with tinnitus, damage to hearing plays an outstanding role. Animal models show that auditory deprivation due to hearing damage causes maladaptive changes in central auditory processing areas, such as increased activity, both spontaneous and synchronised (Eggermont & Roberts, 2004; Schaette, 2014), that is “interpreted” as sound by downstream brain regions. In humans, 75-80% of individuals with tinnitus are also diagnosed with hearing loss using standard pure-tone audiometry (Wallhäusser-Franke et al., 2017). This, however, is likely a conservative lower bound, as hearing damage does not necessarily go along with audiometric hearing loss (Liberman, 2015), which is also evident in individuals with tinnitus (Schaette and McAlpine, 2011; Weisz et al., 2006). Apart from the very mixed findings in humans in support of the hyperactivity/synchrony view (Eggermont & Roberts, 2015), which is also clearly related to the challenge of operationalising putative neural processes using non-invasive methods such as fMRI or M/EEG, there is a clear explanatory gap with regard to the basic assumption that *a majority of individuals with hearing loss do not develop tinnitus* (Lockwood et al., 2002; Tan et al., 2013). Thus, while hearing loss is clearly important, there must be other factors that make some individuals more vulnerable to the development of tinnitus (see e.g., our recent work on prediction engagement; Partyka et al., 2019).

Chronological age has also been associated with the prevalence of tinnitus (Baguley et al., 2013; Biswas et al., 2022; Jarach et al., 2022; for a negative finding however, see Oosterloo et al., 2021). Despite the statistical relationship, the process of ageing is usually not seen as a risk factor on its own, but rather simply related to the deterioration of hearing that goes along with getting older (i.e., age-related hearing loss; presbycusis (World Health Organization, 2021)). However, circumstantial evidence shows that ageing processes might constitute risk factors in their own right. For example, the greatest increase in tinnitus onset is around middle age (<60 years), slightly prior to the greatest increase in onset of moderate to strong hearing loss (Al-Swiahb and Park, 2016). Furthermore, a recent retrospective study shows that tinnitus individuals were 63% more likely to develop early-onset dementia, while controlling for “confounding factors’’, including hearing loss (Cheng et al., 2021).

The aim of the present study is to shed new light on the relationship between (chronological) age, hearing loss and tinnitus. In a first exploratory step, we used data from our ongoing online study *<u>Wie hört Salzburg?</u>*. We then pooled publicly available data from the *<u>National Health and Nutrition Examination Survey</u>* (NHANES, National Center for Health Statistics (NCHS)). Using logistic regressions to predict the presence of tinnitus, we sought to establish ageing as a risk factor. The NHANES data indicated that ageing could make individuals more vulnerable to developing tinnitus when hearing loss is present. Our study underscores the importance of understanding the biological ageing processes that predispose people to tinnitus. A working hypothesis for future studies that synthesises tinnitus with research on hearing loss and cognitive decline will be outlined in the discussion.

## Methods

### Study 1

#### Design and participants

We initially focused on our *Wie hört Salzburg?* online study of hearing epidemiology in the county of Salzburg (Austria). The study included demographic information as well as questionnaires about tinnitus (German short version of Tinnitus Questionnaire, Mini-TQ (Goebel and Hiller, 1992)) and hearing characteristics (German version of the Speech, Spatial and Qualities of Hearing Scale, SSQ (Kiessling et al., 2012)) and an online hearing test (Shoebox, Ottawa, Canada). Participants were instructed to perform the hearing test without any hearing aids. We advertised the study via social media, at the university and by directly contacting older people. We included participants ages 18-80 and excluded individuals with missing information. With this sample, we aimed first for an evaluation of the question whether age per se is a risk factor for tinnitus development.

#### Definition of variables

For our analyses, we focused on variables assessing the hearing abilities and tinnitus conditions of individual participants. The online hearing test yielded a hearing ability score in percentage, which was computed automatically by the provider. Tinnitus was assessed by asking whether participants ever experienced ringing in the ears for a longer period of time. Further, tinnitus distress was investigated using the TQ.

#### Limitations

With respect to the study question, this sample has several limitations. First, age was not uniformly distributed, with fewer participants in the 30-40 age range. Second, the hearing test that we used was a commercial product sold as a screening tool. Results are correlated with the outcomes of pure-tone audiometry but are not suitable as a substitute. Third, we did not differentiate between past, acute or chronic tinnitus in the questionnaire. However, the distribution of hearing problems in our sample matched the results of the World Report on Hearing (World Health Organization, 2021) relatively well. We therefore used the *<u>Wie hört Salzburg?</u>* dataset for exploratory analyses that motivated the analysis of additional data, thus overcoming the aforementioned limitations.

### Study 2

#### Design and participants

For a more representative sample, we included publicly available data from the *<u>National Health and Nutrition Examination Survey</u>* (NHANES, National Center for Health Statistics (NCHS)). Since 1999, the NHANES study has consisted of annual measurements, with approximately 5000 participants added to the database each year. Along with several other types of data, audiometric information was gathered. Individuals had objective pure-tone audiometry and were asked about the presence of tinnitus, the frequency of its occurrence and the level of distress it caused. We merged data from the years 2001 to 2017, including participants up to age 75 with either no tinnitus at all or with chronic tinnitus, which resulted in a total sample size of 16008 individuals.

#### Definition of variables

Hearing was objectively measured via pure-tone audiometry, including seven frequencies from 500 Hz to 8 kHz. For our analyses, we calculated the individual mean hearing ability, averaged across both ears, based on 500, 1000, 2000 and 4000 Hz, which is a common approach for averaging results of pure-tone audiometry (i.e., PTA-4, see for example Lin et al., 2011; Ozdek et al., 2010). Tinnitus occurrence was investigated with the question, “In the past 12 months, have you ever had ringing, roaring, or buzzing in your ears?”. In the case of an affirmative response, participants were asked, “How often did this happen? Would you say…”. Potential answers were, “almost always”, “at least once a day”, “at least once a week”, “at least once a month” and “less frequently than once a month”. For our analyses, we included only participants with either no tinnitus or with tinnitus that occurred “almost always”. Therefore, we targeted chronic tinnitus alone and aimed for the greatest dissimilarity between groups in terms of experiencing tinnitus.

#### Statistical analysis

For both analyses, we used logistic regressions (family=binomial) implemented in R (R Core Team, 2022). Both models aimed to predict tinnitus based on the age and the mean hearing ability of both ears. Models were calculated with and without interaction terms for the predictors age and hearing loss. For the age variable, we split the data by median into two groups, resulting in a cut-off age of 55 years in the first sample and a cut-off age of 34 years in the NHANES sample. In addition, we calculated the odds ratio for the NHANES sample.

#### Role of the funding source

The funders had no role in the study design, data collection, data analysis, data interpretation, writing, or interpretation of the manuscript, nor in the decision to submit it for publication.

## Results

Our first sample consisted of 2249 inhabitants of Salzburg county (1147 female, 4 other, *median=55* years, *mad=13*.*34*). 695 participants indicated that they have experienced tinnitus. Tinnitus was detectable throughout all ages, and age was positively correlated (*r*(2247)=.499, *p*<.001) with mean hearing loss using Spearman’s rank correlation (Figure 1a). Further analyses using logistic regressions emphasised hearing loss as a significant predictor of tinnitus development (*b*=0.017, *p*<.001). However, older age was also a significant predictor (*b*=-0.538, *p*<.001) for tinnitus development – regardless of the extent of hearing loss. A second model, testing a potential interaction between age and hearing loss, yielded no significant effect (*b*=-0.004, *p*=.403) (Figure 1b). These first, exploratory analyses suggested that age per se could be a risk factor for tinnitus. To follow up on these findings in more detail, we used the same methods on the more representative sample of the NHANES study.

**Fig 1:**
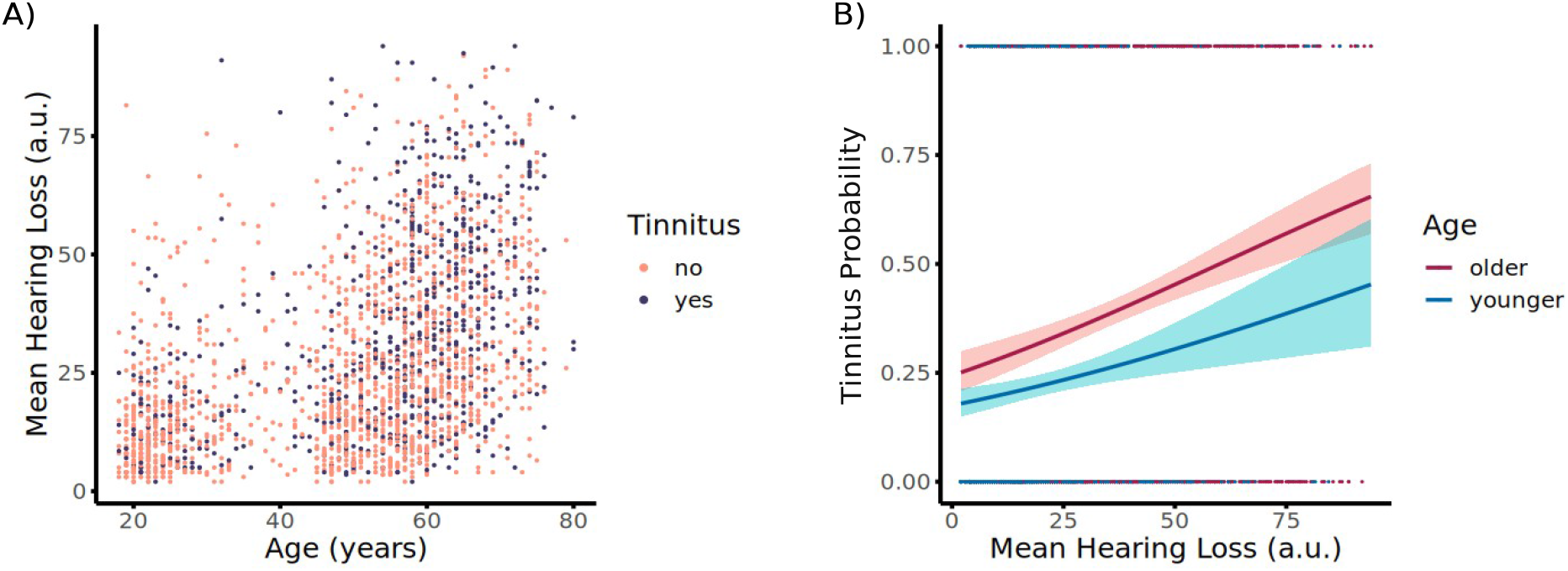
Descriptives and logistic regression model of the Salzburg dataset. **A)** Distribution of tinnitus over age and mean hearing loss. **B)** Age characterization was based on the median age of 55 years. The results showed enhanced tinnitus probability in older individuals compared to younger people over mean-hearing-loss scores.

In this second sample, we included 16008 individuals with an age range between 12 and 75 years (8044 female, *median*=34 years, *mad*=25.20). 1017 participants experienced chronic tinnitus. As figure 2a indicates, the sample is more evenly distributed over age, and age was again correlated with hearing loss using Spearman’s rank correlation (*r*(16006)=.621, *p*<.001). For a descriptive depiction of the sample, we computed the proportion of tinnitus and hearing loss over age groups (Figure 2b). Hearing loss was defined as an average hearing ability over 25 dB. Tinnitus and hearing loss both increase with age, but to different extents. Whereas hearing loss rises rapidly with age, tinnitus peaks in the youngest age group, and then drops between ages 25 and 45. In the older age groups, the tinnitus proportion rises again, but to a much lesser extent than hearing loss. This also highlights that while many elderly people have hearing loss, not all of them also have tinnitus.

**Fig 2:**
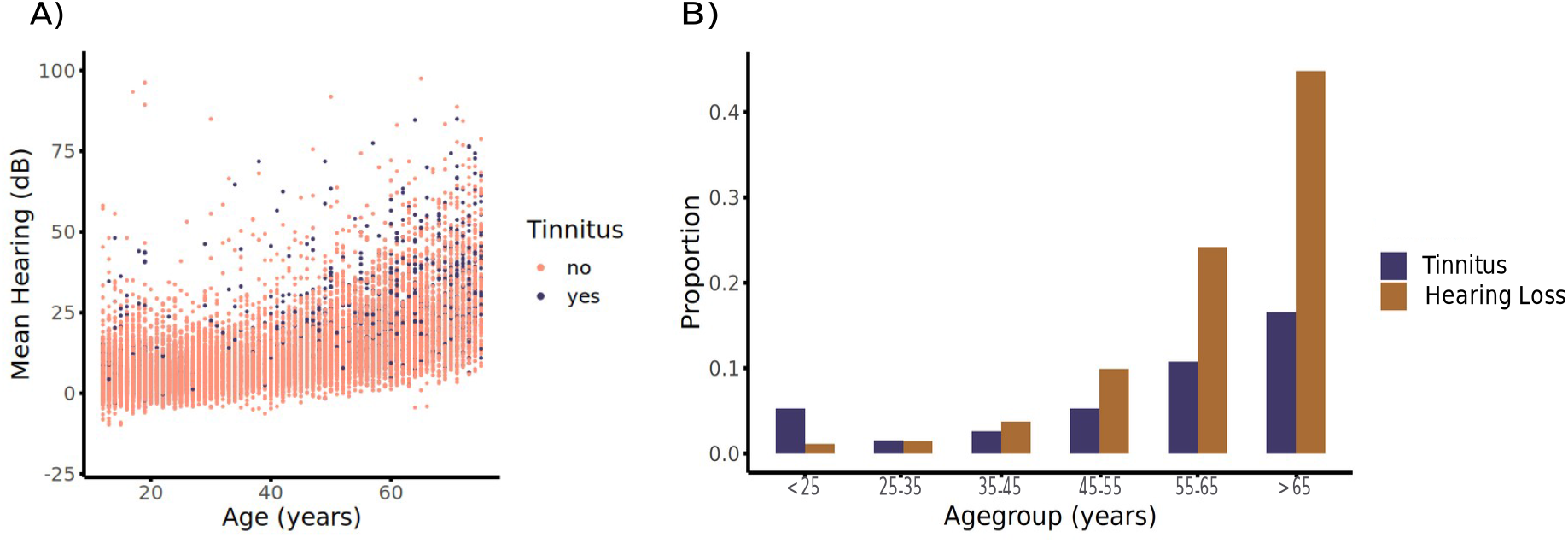
Descriptives of the NHANES dataset. **A)** Distribution of tinnitus over age and mean-hearing threshold. **E)** Proportion of tinnitus and hearing loss in different age groups.

In a simple regression model without interaction term, hearing loss was identified as a significant predictor of tinnitus (*b*=0.057, *p*<.001), whereas age was not significant (*b*=0.058, *p*=.469). However, a second model including a potential interaction term yielded significant effects for hearing loss (*b*=0.061, *p*<.001) and age (*b*=0.352, *p*=.001), as well as an significant interaction effect between the two predictors (*b*=-0.027, *p*<.001). Comparing the two models using a Likelihood Ratio Test showed that the more complex interaction model was more superior in capturing the data as compared to the simpler model (*deviance*=15.1, *p*<.001). The interaction effect suggests that hearing loss predicts the probability to report tinnitus, however that this relationship is different for the two age groups. Indeed Figure 3 clearly shows that the probability to report tinnitus is similar for younger and older individuals when hearing loss is weak. However the difference between the groups grows as hearing loss becomes more pronounced, with older individuals being more likely to report tinnitus.

**Fig 3:**
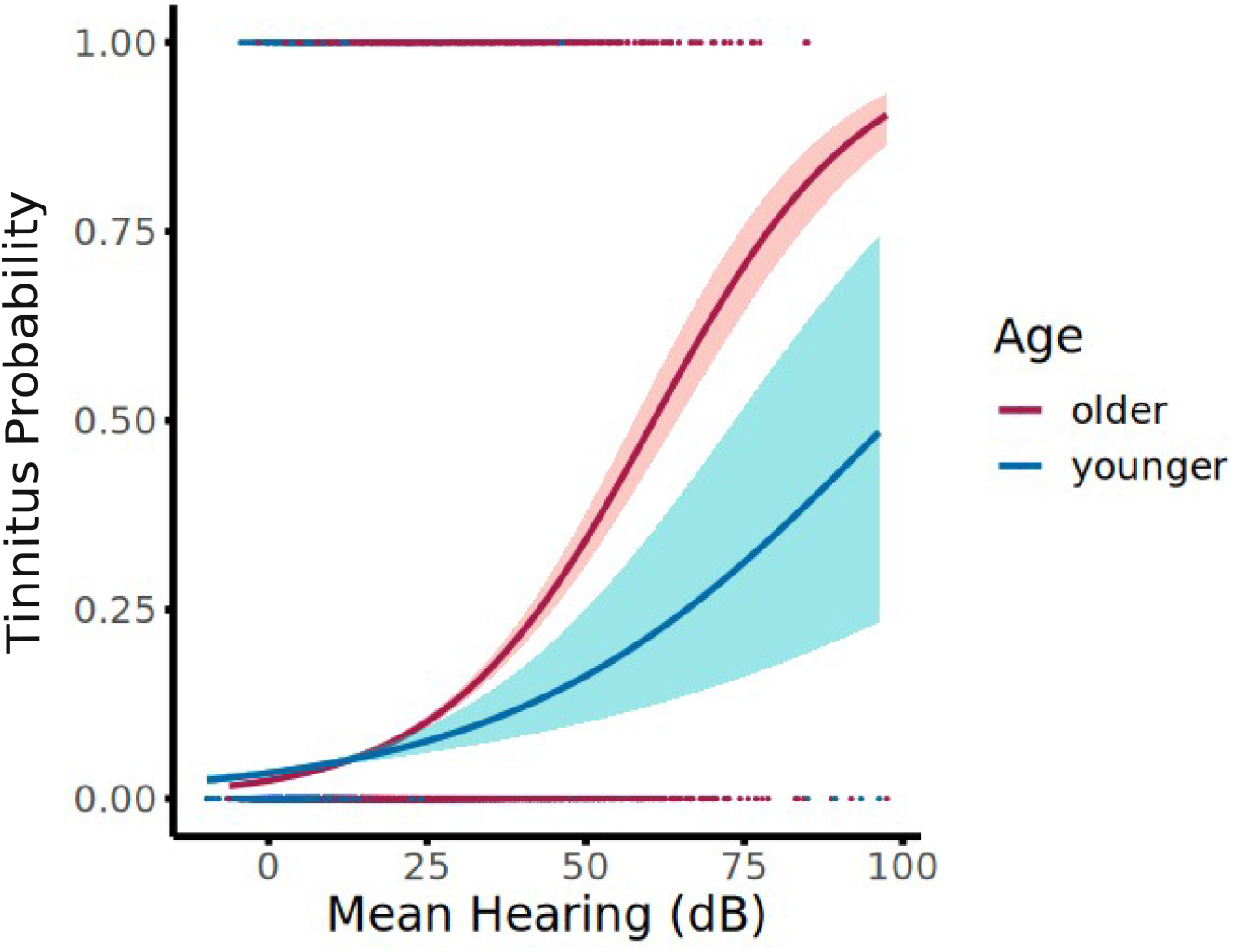
Logistic regression model of the NHANES dataset. Age characterization was based on the median age of 34 years. The results showed enhanced tinnitus probability in older individuals compared to younger people over mean-hearing-loss scores.

To follow up this interaction in more quantitative terms, we calculated the odds ratio (OR) based on the regression coefficients of the second model. For an increase of one-unit in hearing loss (here dB of the pure-tone audiometry), the OR to report for younger people is 1.03 and for older people 1.06. This means, increasing hearing loss by one decibel, enhances the odds of tinnitus by 3% in younger people, whereas the odds for older people increases by 6%. Therefore, for the same amount of hearing loss increase, the odds of older people to report tinnitus are much higher as compared to younger individuals.

## Discussion

The increased prevalence of tinnitus in older age is mostly interpreted in the context of hearing loss (presbycusis), for which mechanistic models also exist that are supported by animal studies. The main motivation of the current study was to understand whether ageing itself poses a risk factor for developing tinnitus. For this purpose, we used two separate study samples to quantify and statistically model the relationship between (chronological) age, hearing loss and tinnitus. The first, exploratory analyses, using an online study of the local Salzburg population, suggested that age per se acts as a risk factor for tinnitus, even when controlling for hearing status. To follow up on these results, we performed the same analysis on a larger sample of participants from the NHANES database (National Center for Health Statistics, (NCHS)). Critically, the NHANES data also provides more precise pure-tone audiometry data. Using this better controlled data set, we were able to find an interaction effect between age and hearing loss. The interaction suggests that the risk to report tinnitus increases with hearing loss, but different for the age groups: estimation of OR clearly showed much higher values in the older as compared to the younger group per unit increase of hearing loss (1.06 vs. 1.03 per dB mean hearing loss increase). Given the diverse nature of the two studies – each with its own purpose – the overall corroborating evidence makes a strong case for viewing ageing (or some other latent biological processes) as a tinnitus risk factor. This work strongly expands previous findings (Schaette and McAlpine, 2011; Wallhäusser-Franke et al., 2017) that focused on hearing loss as a major risk factor for developing tinnitus. Our findings suggest that current models need to be complemented to explain the impact of (chronological) age and hearing loss on developing tinnitus. This likely requires great research efforts into biological ageing processes that make individuals more vulnerable to developing tinnitus.

### Strengths and limitations

A strength of the study design was the inclusion of a highly representative sample in addition to the study we conducted. In the NHANES study, tinnitus was measured using highly differentiating questions to better identify individuals with chronic tinnitus. Additionally, hearing ability was assessed using the “gold standard” of pure-tone audiometry. Another strength of the study was the comparison of two different samples, both highlighting the importance to consider age as a predictor, despite major differences in the two datasets. Interpretation of the Austrian dataset was limited but strengthened the effects, since results derived from it point into a similar direction as the NHANES study. Overall, this data cannot give insights into the exact underlying ageing mechanisms that drive our findings. The present study, however, opens new avenues to guide more mechanistically oriented research and to promote urgently needed conceptual advancements in understanding tinnitus.

### Future directions

#### Towards an integrated understanding of tinnitus, hearing loss and biological ageing

Altogether, our data provides strong support that ageing per se is a risk factor for tinnitus. However, the underlying mechanisms remain unclear. In order for the field to progress, the current study emphasises the necessity of a different line of tinnitus research that seriously considers biological ageing processes and integrates them into existing models. One direction could be to focus on age-related changes in the cochlea (Eggermont & Roberts, 2015). Another line of research could build upon the established relationship between hearing loss and cognitive decline (Lin et al., 2011). While the exact mechanisms are not known (Griffiths et al., 2020), neural degeneration has been reported in individuals with hearing loss, especially in the hippocampus (Xu et al., 2019). Thus, we suggest that hearing damage promotes acceleration of brain ageing (Cole et al., 2019) and that this is the *actual factor* that makes tinnitus more likely. This working hypothesis generates several tangible predictions. For example, it would predict a statistical relationship between tinnitus and cognitive decline, a claim for which some tentative evidence exists (Jafari et al., 2019). Longitudinally, individuals with advanced brain age should be more prone to developing tinnitus. Another, perhaps more-provocative, prediction would be that tinnitus itself is an indicator of advanced or accelerated brain ageing. Conducting large-scale studies using established MRI- or electrophysiology-based measures of brain age (Cole et al., 2019; Engemann et al., 2022) would be invaluable in pursuing this hypothesis.

#### Concluding remarks

Overall, our results are striking testimony in support of recognizing ageing per se as a tinnitus risk factor – rather than solely as an “enabler” of hearing loss, as previously assumed. Since chronological age, as used in the present study, is only a proxy for some latent causes, our work calls for urgent research efforts to be dedicated to linking biological ageing processes, hearing loss and tinnitus. A conceptual integration is necessary, and we have advanced a novel working hypothesis that integrates the various interrelated domains (i.e., hearing loss – tinnitus, hearing loss – neural / cognitive decline, ageing – tinnitus) from a brain ageing viewpoint. Overall, we aim to motivate a series of studies that could significantly advance our understanding and therefore promote innovative treatment and prevention approaches.

## Data Availability

The National Health and Nutrition Examination Survey is a public open database. Data from our online study Wie hoert Salzburg are available to any researcher. The corresponding author of the present study can be contacted to request the data used in the present analysis.

## Supplementary

Summary of the simple and complex model of the NHANES data.

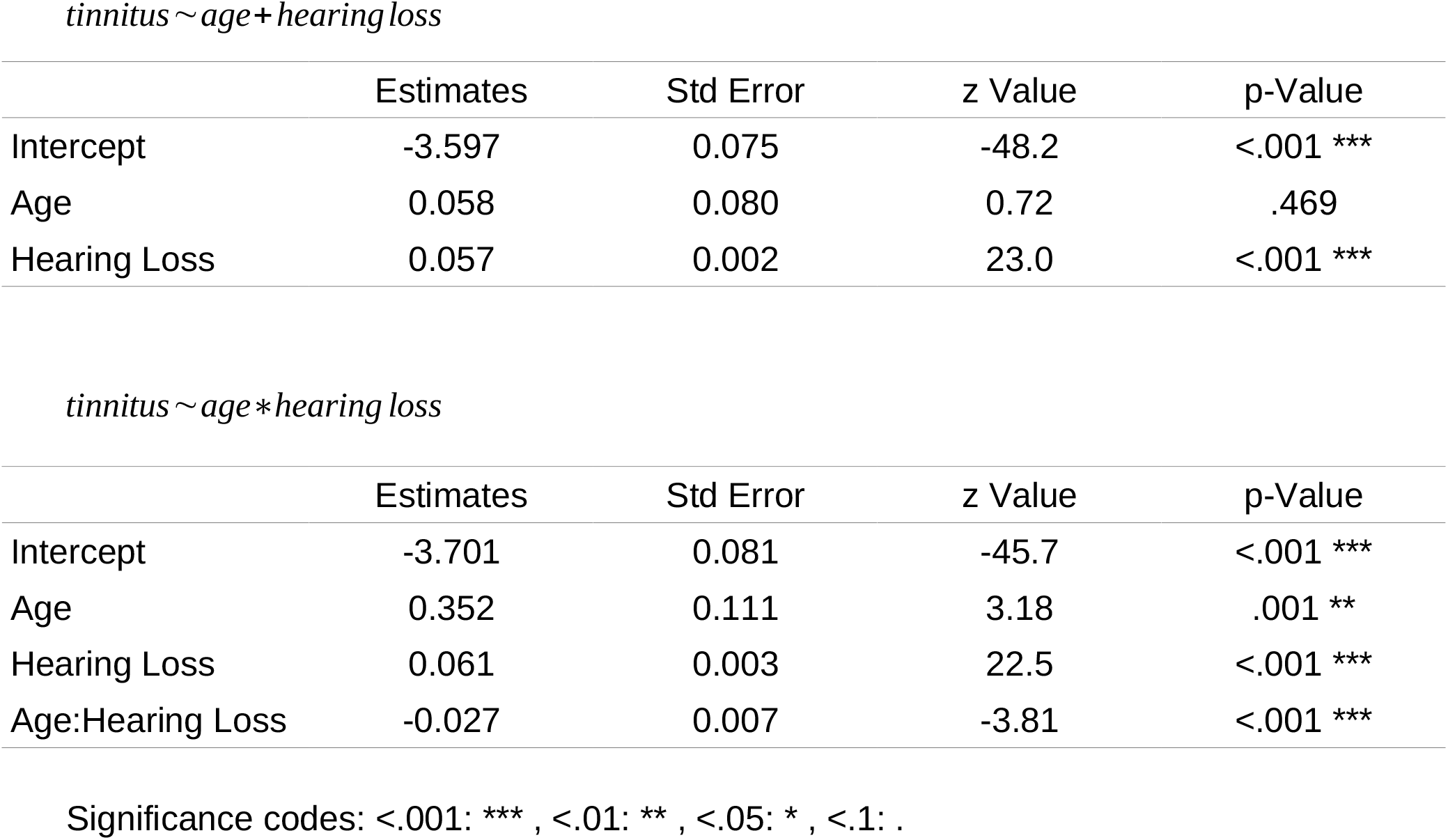

|  | Estimates | Std Error | z Value | p-Value |
| --- | --- | --- | --- | --- |
| Intercept | -3.597 | 0.075 | -48.2 | <.001 *** |
| Age | 0.058 | 0.080 | 0.72 | .469 |
| Hearing Loss | 0.057 | 0.002 | 23.0 | <.001 *** |

*tinnitus ~ age \* hearing loss*
|  | Estimates | Std Error | z Value | p-Value |
| --- | --- | --- | --- | --- |
| Intercept | -3.701 | 0.081 | -45.7 | <.001 *** |
| Age | 0.352 | 0.111 | 3.18 | .001 ** |
| Hearing Loss | 0.061 | 0.003 | 22.5 | <.001 *** |
| Age:Hearing Loss | -0.027 | 0.007 | -3.81 | <.001 *** |

Significance codes: <.001: \*\*\* , <.01: \*\* , <.05: \* , <.1: .

## Notes

### Competing Interest Statement

The authors have declared no competing interest.

### Funding Statement

This study was funded by the Austrian Research Promotion Agency (FFG; BRIDGE 1 project "SmartCIs",871232) and Land Salzburg ("Hidden Hearing Loss", 20204-WISS/225/288/4-2021).

### Author Declarations

The ethics committee of the University of Salzburg (EK-GZ: 22/2016 with Addenda) gave approval for this work.

